# Classifying polyneuropathy and myopathy patients on Electronic Health Records

**DOI:** 10.64898/2025.12.11.25342051

**Authors:** Md Shamim Ahmed, Nicolai Dinh Khang Truong, Elisabeth Nyoungui, Jiawei Zhao, Hanna Wedemeyer, Rudolf Mayer, Verena Schuster, Jana Zschüntzsch, Richard Röttger

## Abstract

**Background:** Rare neuromuscular diseases such as polyneuropathy (PN) and myopathy (MY) often share symptomatic characteristics, leading to diagnostic challenges and delays. Machine learning applied to routine care data of electronic health records (EHRs) offers the potential for accelerating accurate diagnosis.

**Objective:** To develop and evaluate machine learning models to distinguish between patients with PN and MY using EHR data, as a step toward tools that could support improved diagnostic processes.

**Methods:** We analyzed EHR data from 2,181 patients (1,853 PN, 328 MY) provided by the Medical Data Integration Center of the University of Göttingen. The features were curated according to the recommendations of the physicians, the literature, and statistical analysis. We implemented Logistic Regression, Random Forest, and XGBoost models, optimized with Grid Search, and addressed class imbalance using SMOTE.

**Results:** Random Forest and XGBoost models achieved the best performance with F1 Macro scores of 0.82-0.84 and AUC-ROC scores of 0.92-0.93 when trained on demographic data, feature-engineered variables, laboratory test results, and ICD-10 codes. Patient age emerged as a significant predictive factor, with MY patients typically diagnosed at younger ages (mean=51.39) than PN patients (mean=67.11).

**Conclusion:** Machine learning models can effectively differentiate between PN and MY patients using EHR data with low data depth, potentially accelerating diagnostic processes for these rare neuromuscular diseases.

**Availability and implementation:** https://gitlab.sdu.dk/screen4care/classifying-pn-and-my

## Introduction

Rare diseases (RDs) affect small population segments, with approximately 5,000 to 8,000 identified worldwide [1, 2]. Patients with RD typically endure long diagnostic journeys that average seven years to a definitive diagnosis, resulting in substantial socioeconomic burdens [3, 4, 5]. Diagnostic delays stem from overlapped symptomatology between RDs and common conditions, heterogeneous presentations, and limited clinician familiarity. Although genetic testing provides a definitive diagnosis when appropriately targeted, resource constraints require selective application guided by strong clinical suspicion [6, 7, 8]. Recent studies have used electronic health records (EHRs) and machine learning (ML) to identify high-risk RD patients [9, 10, 11].

EHRs generally contain structured and unstructured longitudinal medical data. These data often include demographic data of the patient, medical history, laboratory test results, medication, vital parameters, and clinical notes [12, 13]. However, EHRs present inherent limitations, including data missingness, heterogeneous structures, and collection bias that compromise ML model performance [14, 15].

In Germany, the Medical Informatics Initiative has mitigated some of these issues by installing Data Integration Centers (DICs) in all university hospitals. DICs harmonise core EHR elements—ICD-10-GM codes, procedures, laboratory values, medications, and selected imaging results—within a common interoperability layer, creating a higher-quality substrate for research use [16].

Leveraging this infrastructure, we adopt a syndromic-AI approach: rather than hunt for single (ultra)-rare entities, the model learns symptom clusters that characterise broader disease families. We target two neuromuscular umbrellas: polyneuropathy (PN) and myopathy (MY). Both can manifest with muscle weakness, pain, and multisystem involvement. Yet, they demand different diagnostic work-ups (e.g., nerve-conduction studies vs. muscle biopsy) and divergent treatments (immunotherapy for chronic inflammatory demyelinating PN vs. genetic counselling and targeted rehabilitation in MY). Their clinical overlap and low prevalence, therefore, make PN and MY separation an ideal testbed for ML applied to real-world DIC data.

Therefore, our primary aim was to develop and thoroughly test a classifier system on routine care EHR data to distinguish between PN and MY, which, if successful, could significantly shorten time-to-diagnosis for neuromuscular RD patients and demonstrate the broader potential of DIC-based syndromic AI. To that end, we apply Logistic Regression (LR) [17], Random Forest (RF) [18] and eXtreme Gradient Boosting (XGBoost) [19] to classify PN and MY using available modalities such as demographics, ICD-10-GM codes, and laboratory results. The second aim was to identify the most informative and useful clinical features for classification, which would also allow to identify potential gaps in the current documentation of patients in routine care data.

## Methods

### Dataset

The study cohort was extracted from the University Medicine Göttingen (UMG) DIC, which harmonises routinely EHRs in compliance with the German Medical Informatics Initiative. After approval by the UMG Ethics Committee (ref. 3/3/24 V.01, 06.02.2024) and completion of a data protection impact assessment, a data-use agreement permitted de-identified data processing. Raw data export (01 Jan 2013 - 31 Dec 2023) contained 2,494 adults (≥ 18 years) with at least one inpatient or outpatient encounter coded for PN or MY. Available modalities comprised:

- Demographics (age, sex)
- ICD-10-GM diagnosis codes
- Laboratory test results with time stamps

The patients were admitted to the hospital for a variety of reasons. A total of 4,172 unique clinical features were identified, one of which has 2,910 ICD-10-GM codes and 1,262 laboratory test results. The frequency of diagnosis encounters per patient ranged from 1 to 13, with a median of 1. Inter-visit intervals varied between 0 days and 54 months (median = 114 days; IQR = 73-273 days).

The patients are labeled as PN or MY if they possessed at least one ICD-10-GM diagnostic code within the range of G60-G64 or G70-G73, respectively (Table S1 and S2). Three exclusion criteria were applied to allow the models to classify PN and MY (Figure 1). First, patients with concomitant PN and MY codes (n = 225) were excluded. Second, patients with missing PN or MY codes (n = 23) were removed from the analysis. Third, patients without recorded codes (n = 65) were excluded, as these cases likely represented one-day administrative visits that would not contribute meaningful clinical information to the analysis.

**Fig. 1:**
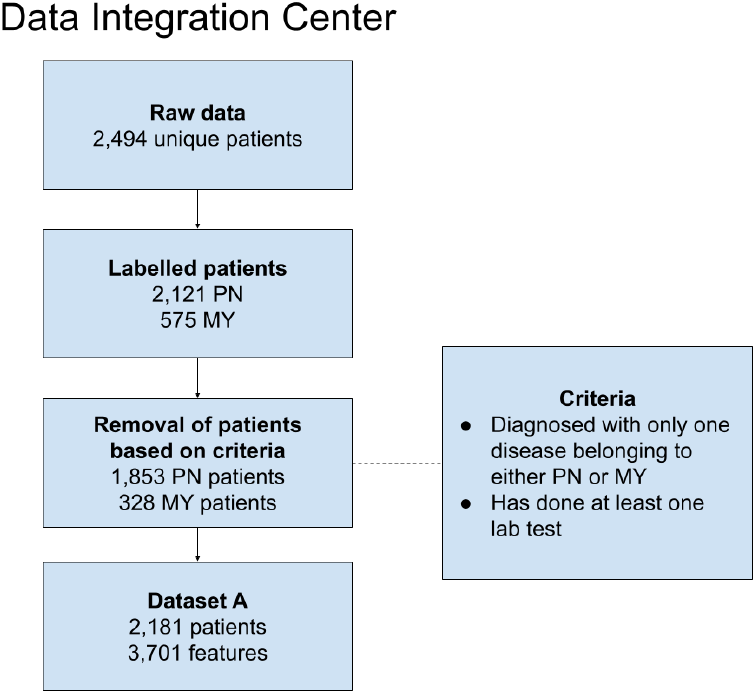
A summary of the data preprocessing on the EHRs of PN and MY patients. PN and MY respectively stand for polyneuropathy and myopathy.

The resulting analytic dataset comprised 2,181 unique patients (PN = 1,853; MY = 328) and 3,701 features. We define this dataset as Dataset A (See Table 1). Supplementary Tables S1, S2, S3, and S4 detail diagnostic code distributions before and after filtering. To preserve demographic balance, the dataset was randomly stratified into 80% / 20% on age group and sex, yielding a training set of 1,744 patients and a held-out test set of 437 patients. The source code is available at https://gitlab.sdu.dk/screen4care/classifying-pn-and-my.

**Table 1.** We created six datasets based on our analyses. In particular, dataset F has undergone dimensionality reduction using Principal Component Analysis based on dataset E. Here, we trained on the first 11 principal components.

| Name | Demographics (De) | Feature Engineering (Fe) | Laboratory Results (La) | ICD-10-GM Codes (Ic) | Total Features |
| --- | --- | --- | --- | --- | --- |
| Dataset A (De+La+Ic) | 3 | 0 | 1,126 | 2,572 | 3,701 |
| Dataset B (De) | 2 | 0 | 0 | 0 | 2 |
| Dataset C (De+Fe) | 3 | 70 | 0 | 0 | 73 |
| Dataset D (De+Fe+La) | 3 | 3 | 40 | 0 | 46 |
| Dataset E (De+Fe+La+Ic) | 3 | 3 | 40 | 30 | 76 |
| Dataset F | N/A | N/A | N/A | N/A | 11 |

#### Data exploration & Normalization

As one of the first steps, we investigated the distributions of the variables provided, particularly the numerical lab values. Here, we observed that highly skewed distributions with extreme values were prevalent, which consequently do not follow the Gaussian distribution. Some machine models, particularly linear models, require normal or at least near-normal distributed features and consequently would benefit from a normalization of those features.

Hence, we conducted a normality test, specifically the Shapiro-Wilk Normality Test [20] on all numerical features, excluding categorical and binary features. Those that failed the normality test were log-scaled. We subsequently tested our models with two versions of the dataset: (a) we only normalized the numerical features with a min-max normalization, regardless of their distribution, and (b) we log-scaled non-normal distributed features, while the normally distributed features were min-max normalized. This step was done to exclude the potential influence of the normalization technique on the model performance. Please refer to Figure 2 for an overview of the data preparation workflow.

**Fig. 2:**
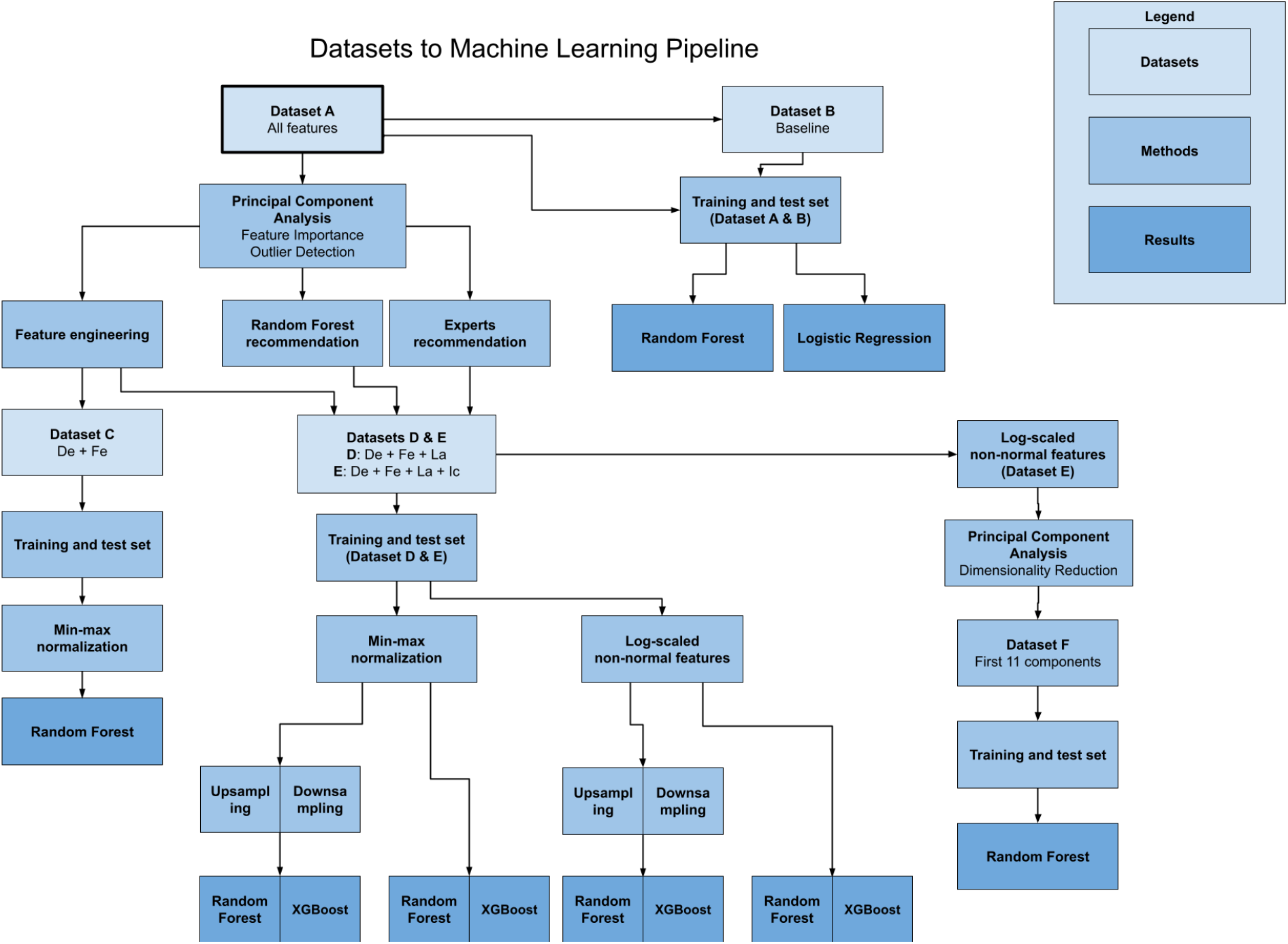
Flowchart of methods. After removing patients who did not fulfill the criteria, as seen in Figure 1, we did feature selection for different purposes. First, we trained a Random Forest with all features (Dataset A) and then trained a Logistic Regression model and a Random Forest on the baseline dataset of age and gender (Dataset B). Thereafter, we created three datasets (Dataset C, D, and E) based on three main methods: feature importance derived by Random Forest, expert-driven feature selection, and feature engineering. *De* stands for demographics; *Fe* for engineered features; *La* for laboratory results; *Ic* for ICD-10-GM codes. The Random Forest recommendation is trained on 3,701 features, which provided a ranked list of important features. Most models are trained with up-sampled or down-sampled training sets. We also log-scaled dataset E’s features, which did not pass the normality test, to perform Principal Component Analysis. Here, we have identified important features and interesting outliers.

### Feature Selection and engineering

Feature selection and engineering are the crucial steps in dataset pre-processing to efficiently classify between MY and PN patients. This involves identifying the most relevant features for the given problem to minimize noise and computational complexity. Using Dataset A, we performed several analyses to identify meaningful features, resulting in five datasets. Figure 2 shows an overview of the process of creating these datasets, which are later applied to ML models.

#### Expert-driven features selection

Expert-driven feature selection was conducted through a systematic collaboration with neuromuscular disease specialists at UMG, employing established knowledge for biomarker identification in rare neuromuscular disease research. Laboratory parameters were selected based on their clinical significance across primary etiological pathways of neuromuscular disorders: inflammatory, metabolic, toxic, and hereditary mechanisms.

Creatine kinase (CK) serves as the primary indicator of muscle integrity [21]. In MYs, CK elevations typically demonstrate substantial increases, with inflammatory MY showing 10-to 50-fold increases above normal reference ranges. Muscular dystrophies and some metabolic MYs show marked early-stage CK elevations that decline progressively as muscle mass decreases [22]. Reference ranges demonstrate significant demographic variability: Caucasian males typically have values 200 U/L, while African-American men can have normal values up to 1,000 U/L, with generally lower values in women of all ethnicities [23, 24]. CK elevation also occurs in PNs and motoneuronal diseases, although typically to a lesser degree than in primary myopathies [25, 26]. Alanine aminotransferase (ALT) and aspartate aminotransferase (AST) function as dual-purpose biomarkers, reflecting both hepatic function and muscle damage, providing complementary information to CK levels [27, 28, 29, 30].

Several additional laboratory markers provide information on the etiology of the underlying PN or MY. Some of these markers are also potentially collected as part of routine diagnostics for other diseases, whereas others are specific to the respective specialised diagnostics.

For PN etiology, high-yield screening includes blood glucose and hemoglobin A1C (HbA1c) for the diabetic neuropathy detection [31, 32], the most common cause. Additional markers include serum B12 with metabolites (methylmalonic acid with or without homocysteine), serum protein immunofixation electrophoresis (detecting paraproteinemic neuropathies) [33], values for kidney (glomerular filtration rate (GFR)) [34, 35] or liver dysfunction, endocrinological (thyroid-stimulating hormone (TSH P and TSH rP). Many PN and MY diseases have autoimmune origins. Immunological markers, such as immunoglobulin A, G, and M, (IgA S, IgG S, and IgM S), assess immune system activity and potential autoimmune triggers [36].

MY are characterized by a higher prevalence of genetic etiologies, reflected in fewer routine laboratory abnormalities beyond CK and transaminases. Specific markers like Mi-2 alpha, Mi-2 beta, and TIF1 gamma are directly related to autoimmune myositis and provide a targeted approach for diagnosing subtypes of MY [37].

Comprehensive inflammatory assessment encompasses infectious serological markers (borreliosis, hepatitis, or HIV), erythrocyte sedimentation rate, C-reactive protein, or rheumatological parameters (ANA, ANCA, or rheumatoid factor), which may indicate inflammatory, autoimmune causes or complications of PN and MY [38]. Hemoglobin (HB) and leukocyte count (Leuko) offer valuable information regarding hematological/oncological disease or immune dysfunctions. The addition of electrolytes such as potassium (K P), sodium (Na P), and chloride (Cl poc) helps achieve a proper understanding of the electrolyte imbalance that might exacerbate neuromuscular presentations or give insights into disease etiology [39, 40].

Selected laboratory parameters were integrated into comprehensive biomarker panels (Datasets D and E), ensuring clinical relevance while maintaining statistical power for machine learning-based PN and MY classification.

#### Feature Engineering

The demographics and laboratory test results exhibited significant differences. The longitudinal data were analyzed statistically, considering measures such as mean, median, standard deviation, minimum, and maximum test values. These laboratory test results were dichotomized into normal and abnormal categories to reduce the effect of outliers (that might have clinical significance), dependence on non-uniform value distribution, and make the model more robust. Table 2 summarizes the results of these markers. Patients with MY exhibited elevated abnormal values, such as ALT P, AST P, and CK P, compared to patients with PN, which aligns with the findings of existing works [27, 28, 21]. In contrast, PN patients showed notable differences in Ery, Gluc-P, and HbA1c.

**Table 2.** Summary of the selected laboratory test results, flagged as normal and abnormal for each patient per disease group. The ratio is calculated as the number of abnormal cases divided by the number of normal cases. The highest ratio in each row is marked in bold font.

| Laboratory tests | PN (n = 1853) |  |  | MY (n = 328) |  |  |
| --- | --- | --- | --- | --- | --- | --- |
|  | Normal | Abnormal | Ratio | Normal | Abnormal | Ratio |
| ALT P | 1290 | 199 | 0.15 | 139 | 97 | **0.70** |
| AST P | 1063 | 441 | 0.41 | 146 | 150 | **1.03** |
| CK P | 1041 | 452 | 0.43 | 137 | 170 | **1.24** |
| CRP P | 845 | 938 | **1.11** | 232 | 83 | 0.36 |
| Ery | 929 | 911 | **0.98** | 269 | 56 | 0.21 |
| Gluc-P | 508 | 765 | **1.51** | 69 | 68 | 0.99 |
| HbA1c | 465 | 609 | **1.31** | 35 | 31 | 0.89 |
| K P | 1617 | 212 | **0.13** | 296 | 24 | 0.08 |
| Na P | 1628 | 200 | **0.12** | 309 | 11 | 0.04 |

In addition, we investigated whether the patient’s age and length of hospital stay in days could act as critical predictive variables for distinguishing PN from MY. Figure 3 shows the age distribution and the median aggregated length of hospital stay. We observe that patients diagnosed with MY (mean age = 51.39) are younger at diagnosis compared to patients with PN (mean age = 67.11). Patients with MY (median stay = 2) tend to stay in the hospital for a shorter period compared to patients diagnosed with PN (median stay = 6). The patient’s age can also play an important role in interpreting laboratory test results. For example, older patients have a physiological increase in creatinine and urea levels. On the contrary, albumin and other proteins tend to be lower in patients of older age. [41]

**Fig. 3:**
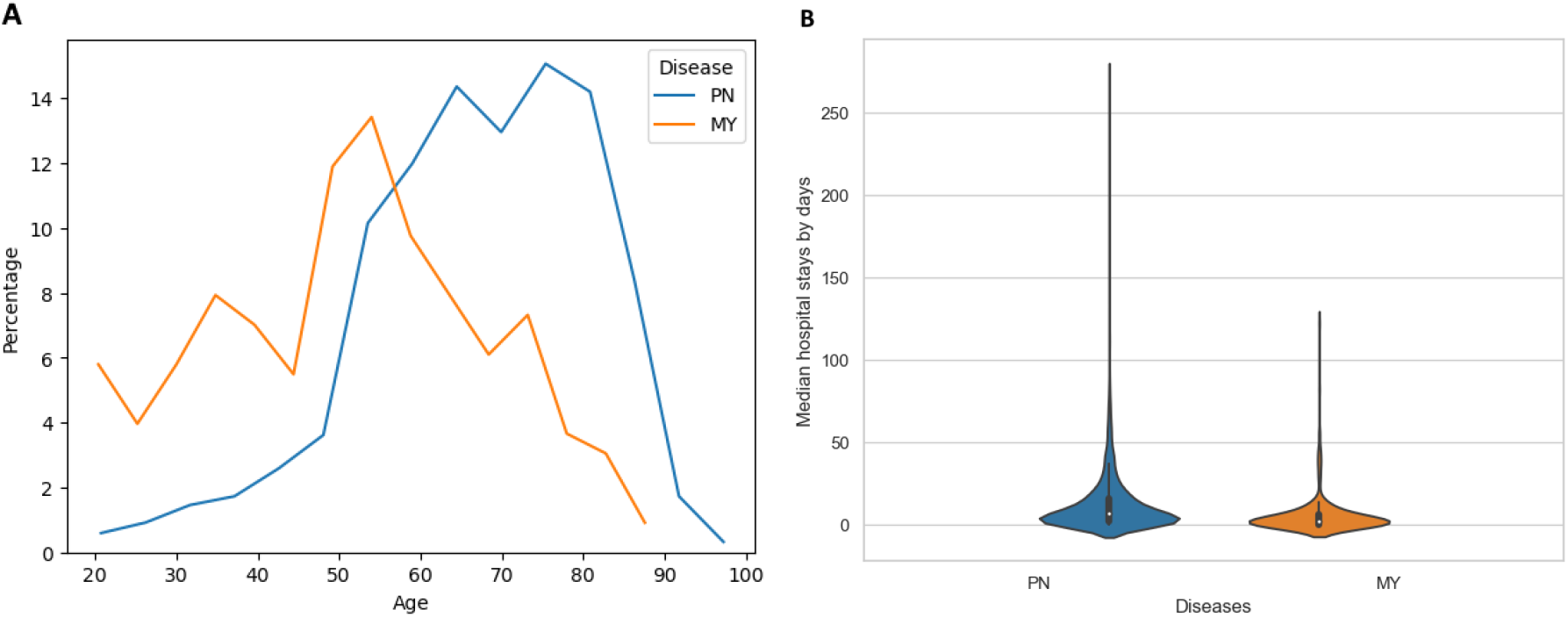
Summary of the age distribution (A) and length of stay at the hospital (B) by diseases. Myopathy (MY) patients are diagnosed earlier in their lives compared to polyneuropathy (PN) patients. Since the patient can visit the hospital multiple times, we have aggregated the length of their stay by taking the median for each patient. PN patients stay longer at the hospital, with a few extreme cases lasting 273 days.

In the statistical analysis of the data, we worked with two main modules: demographic features (such as age, gender, and living status) and laboratory test results. Many patients had the same laboratory tests multiple times, often at different points in their hospital visits. The dataset’s temporal components were analyzed to evaluate the potential for time-series machine learning analysis. To make the best use of the irregular dataset, two approaches were considered:

##### Time Series Analysis

Track how the results of the laboratory tests of each patient changed over time. **Aggregated Analysis**: Summarize the results of the laboratory test for each patient by computing simple statistics (such as mean and median, standard deviation, minimum value, and maximum value) in multiple tests.

Due to the irregular timing of laboratory tests (i.e., they were not always taken at the same intervals for all patients), aggregated analysis proved to be a suitable option, which allowed us to capture important trends without the need for evenly spaced data. Following these approaches, Dataset C was created, which contained demographics and engineered features.

#### Machine Learning-Based Selection

Following the initial data cleaning, we conducted an exploratory phase on all 3,701 available features in Dataset A. This set consists of features that include laboratory test results, patient demographics, and diagnosis codes in the form of ICD-10-GM codes. An ML-based feature selection approach was incorporated by training an RF model to classify PN and MY. This model allowed us to develop a basic predictive model and gain insight into the importance of each feature. From this analysis, we identified the 40 most significant features as depicted in Figure 4.

**Fig. 4:**
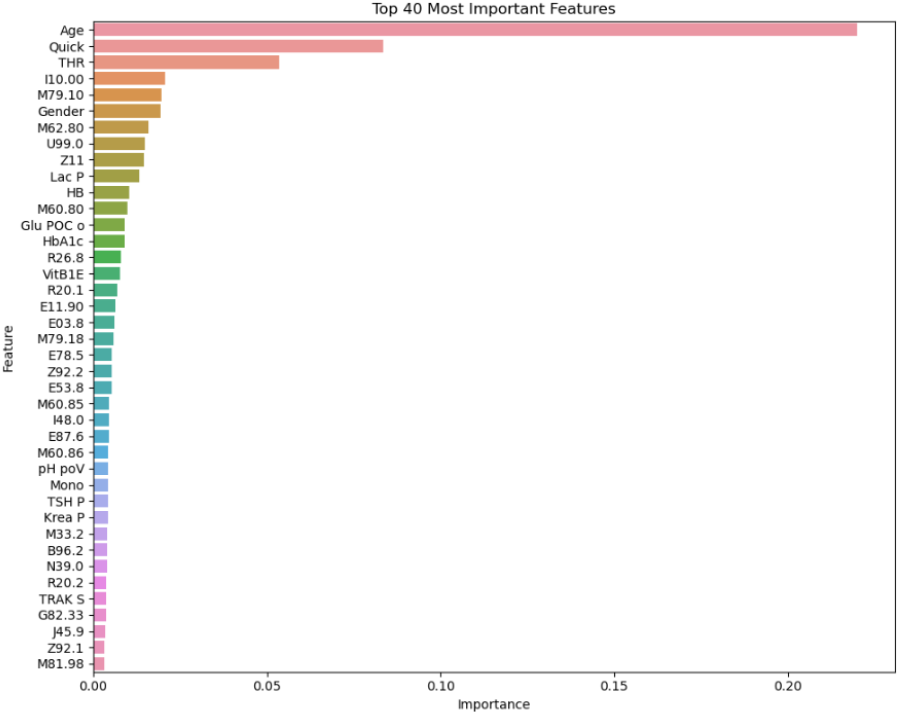
Feature importance using RF using 3,701 features. In particular, the five most important features consist of demographic information (age), laboratory test results (Quick and THR), and German-modified ICD-10 codes (I10.00 and M79.10). This helped select features that could be important for generating datasets.

This basic model, with all its features, yielded some modest performance. Despite the large number of features, we realised that the data had some inherent patterns or directions that could result in a meaningful classification of PN and MY in this phase. Hence, the important features from the initial analysis did contribute to the selection of features for generating Datasets D and E. For example, features such as Age, Gender, M79.10.

#### ICD-10-GM codes

Given the heterogeneity of patient presentations and comorbidities in university hospitals, certain ICD-10-GM codes may be positively correlated with PN or MY diagnoses. As diabetes mellitus is the most prevalent cause of PN in Europe [42], it is reasonable to anticipate a higher prevalence of diabetes mellitus as a comorbid diagnosis in PN patients compared to MY patients. Incorporating ICD-10-GM codes into diagnostic models could enhance the differentiation of PN and MY patients, which leverages the distinct etiological profiles of these conditions.

The most frequent ICD-10-GM codes excluding codes used for diagnosing PN and MY were identified (see Supplementary Tables S5 and S6) and combined those derived from Section 2.2.3. The ICD-10-GM codes related to the examination of irrelevant diseases were excluded. For example, Z11 is used to screen infectious and parasitic diseases. The findings were combined with the ML-based feature selection method, resulting in the formation of Dataset E. The main difference is that Dataset D does not contain any ICD-10-GM codes.

#### Principal Component Analysis

The EHR data that we used has a high number of features. In the scenario of a large number of features, training models on all features could potentially deteriorate the performance, as the majority of these features are not fine-grained and hence may not provide meaningful contributions to the model. This phenomenon is known as the curse of dimensionality [43]. We incorporated Principal Component Analysis (PCA) to reduce the dimensionality of a dataset while preserving maximum possible variance [44]. The first *k* principal components (PCs) are utilized as features for machine learning models. The PCs are determined using the elbow method on the Eigenvalues. In this study, we have used these principal components to form Dataset E (See Table 1).

Additionally, we used the first few principal components to visualize the dataset. The projection of the original, non-normalized dataset, depicted in Supplementary Figure S2. It shows that the dataset suffers from a significant number of outliers, highlighting the importance of log-scaling of the features with highly skewed distributions. The usage of log-normalized features instead resolved the issue of extreme values of some of the lab-values and shows that the two groups are rather convoluted and will be difficult to separate, at least in a linear fashion, as shown in the Figure 5.

**Fig. 5:**
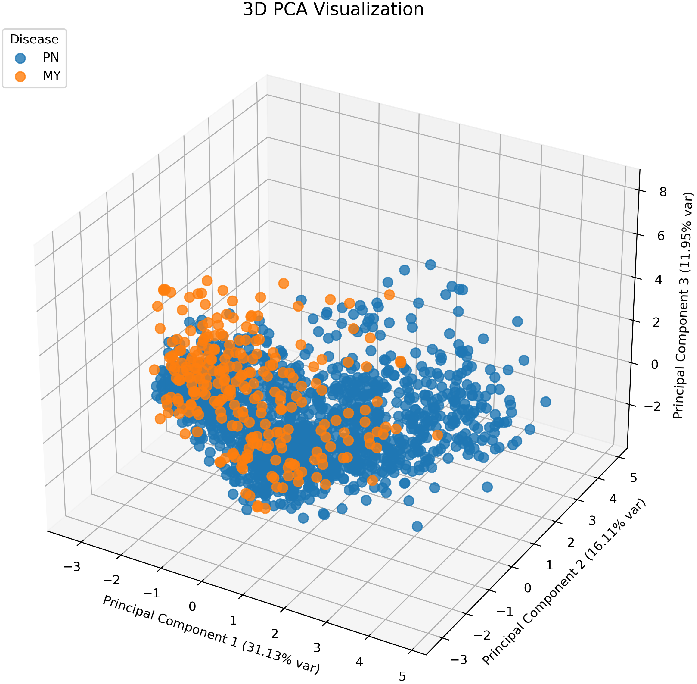
3D Visualization of PCA after log-scaling non-normal features.

### Grid Search

Grid Search is a hyperparameter optimization method that exhaustively combines all available hyperparameters of a model to achieve optimal results. Grid Search was applied to all our models. The cross-validation and selected hyperparameters were applied to achieve the highest mean F1 Macro score. Section 2.5 provides an explanation and overview of the metrics we have selected for evaluation. Supplementary Table S11 shows a list of hyperparameters for each model for Grid Search.

### Machine Learning Approaches on Feature Sets

We incorporated LR, RF, and XGBoost ML algorithms for our experiments. LR is a simple and interpretable statistical method for predicting binary outcomes [17]. RF combines decision trees trained on the sampling of data and sub-sampling of features. After all decision trees are formed, these trees will decide on the predicted class based on average class probability or majority voting. This model is popular in clinical settings [18, 45, 46]. Unlike RF, XGBoost trains trees sequentially, where each successor focuses on correcting errors from the previous tree by training on adjusted weights [19]. Both RF and XGBoost are capable of handling features with unknown values, serving as ideal candidate ensemble models for patients who exhibit a range of symptoms, comorbidities, and have undergone different laboratory tests. RF and XGBoost were applied to most of the datasets as they were capable of classifying diseases given a patient record with missing data.

#### Baseline Models

We selected features ‘age’ and ‘gender’ to train a baseline model, because these are often powerful predictors in clinical settings and therefore can be compared while we build better models. Figure 3 shows the age distribution of patients at diagnosis for PN and MY. Here, we can observe that many MY patients were diagnosed earlier in their lives, while PY patients were generally diagnosed at later ages. The RF model identifies gender as a fairly important feature in Figure 4. Aaberg et al. [47] conducted a study showing that male patients develop diabetic polyneuropathy earlier than females. At the same time, age and gender have been attributed to neuromuscular diseases and often influence the prevalence and progression rate [48, 49]. For example, PY associated with diabetes mellitus, monoclonal gammopathies, and malignancies occurs more frequently at an older age [50]. There are also forms of MY that tend to occur at older ages. While these are mostly acquired, MY forms of genetic facioscapulohumeral muscular dystrophy (FSHD) can also occur in elderly patients [51]. Hence, the models in this study were trained with age and gender as input features to provide a baseline for model performance. These baselines would allow a straightforward comparison to determine whether other complex features significantly improve the performance compared to these models.

LR and RF algorithms were implemented to train models for baseline performance evaluation. LR was applied due to its simplicity and interpretability, while RF may provide insights into non-linear correlation and possible interactions between age and gender.

#### Model Using Demographic and Statistical Features

After aggregating the laboratory test results as mentioned in Section 2.2.2, Dataset C was created, consisting of demographic and descriptive statistics of laboratory test results, to build the model using RF. Demographic information (age, gender, living status) provided an essential background of each patient, whereas summarized laboratory test results provided deeper insights into their health patterns over multiple visits.

#### Expert-Driven Features + Random Forest recommended Features + Statistical Features

As discussed in the previous sections (Figure 2), various feature selection strategies were employed. Ultimately, two distinct feature sets were created by combining expert-driven features, Random Forest-recommended features, and selected statistical features. Dataset D consisted of demographic features, engineered features, and laboratory test results, while Dataset E additionally included selected ICD-10-GM codes.

The demographic features comprising ‘age’ and ‘gender’ were included in these feature sets. To further enrich these datasets, variables such as the number of days the patient stayed in the hospital were included, which could potentially provide insights into disease severity and progression. Two statistical features were also included: **Deviation from Min**: The difference between the patient’s laboratory test result and the lower bound of the normal reference range. **Deviation from Max**: The difference between the result of a laboratory test and the upper bound of the normal reference range.

PCA was applied to address the high dimensionality in Dataset E. A Scree plot (Supplementary Figure S4) showed that the first 11 principal components were sufficient for training the models.

This version of the dataset, comprising 11 principal components was termed as Dataset F. Thereafter, RF and XGBoost models were trained separately on Datasets D, E, and F.

#### Handling of imbalanced data

As the training set is imbalanced, the impact of up-sampling or down-sampling on the training set was assessed. The imbalanced training set had the limitation that the models performed worse while classifying the minority class MY. It is important to note that the test set is not up-sampled or down-sampled to reflect reality. This is later used to evaluate the datasets in Section 3. Firstly, down-sampling was incorporated by randomly downsizing PN patients until the training set was completely balanced. Secondly, up-sampling was incorporated to handle the imbalance in the training set. The up-sampling algorithms, Synthetic Minority Over-sampling technique (SMOTE) and Adaptive Synthetic Sampling (ADASYN), were considered. Both methods generate synthetic samples for the minority class by interpolating between existing samples, effectively increasing the representation of the underrepresented class. While both methods yielded similar results regarding model performance, SMOTE was selected due to its simplicity and common usage in medical datasets [52].

### Model Evaluation Metrics

Distinguishing between PN and MY is crucial due to their symptomatic similarities. While specific diseases falling under the families of PN or MY are important to diagnose, a balanced focus is needed to classify both groups as effectively as possible. Therefore, the following metrics are selected: Accuracy, F1-Macro, Area Under the Curve Precision-Recall (AUC-PR), and AUC-ROC to evaluate the performance of the models. Accuracy measures how well a model predicts overall outcomes.

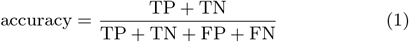

where TP, TN, FP, and FN are the numbers of true positives, true negatives, false positives, and false negatives, respectively. While accuracy is a standard metric for measuring performance, it does not consider class imbalance. Precision measures the accuracy of positive predictions.

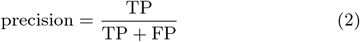

Recall measures how well the model identifies the cases of one class.

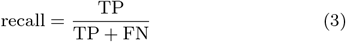

To account for class imbalance, model performance was evaluated using the F1 score. The F1 score, calculated as the harmonic mean of precision and recall, provides a balanced measure of a model’s performance for a single class.

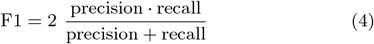

In contrast, the F1 Macro score evaluates a model’s ability to classify across multiple classes to ensure a balanced inter-class evaluation.

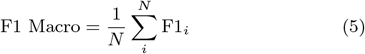

where *N* is the number of classes. Applying the F1 Macro score is important for evaluating a model’s performance across all classes.

AUC-ROC is another popular metric for evaluating performance in binary classification tasks [53]. The ROC curve depicts the trade-off between the True Positive Rate (TPR) and the False Positive Rate (FPR) across various classification thresholds. These two rates are calculated as follows:

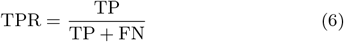

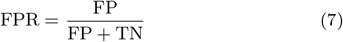

The Area Under the Curve (AUC) summarizes a model’s performance in a single metric:

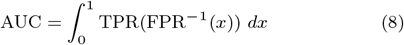

where 0.5 corresponds to random guessing and 1.0 indicates a perfect classifier. Hence, the higher the AUC, the better the model distinguishes the positive and negative classes. Tailored to the PN versus MY classification, the former is designated to be the positive class and the latter to be the negative class. Therefore, the TPR shows how well the model correctly classifies the PN patients, and the FPR shows how much our model misclassifies the MY patients.

However, AUC-ROC assumes there is a class balance. The AUC-PR measures the trade-off between the precision and recall of a positive class. As the datasets are heavily imbalanced in favor of PN, the analysis focused on how well the models classified MY patients.

## Results

### Data exploration using Principal Component Analysis

During data exploration, dimensionality reduction was performed using PCA described in Section 2.2.5 on the raw dataset to identify outliers and meaningful features. The dataset contains some notable statistical outliers (See Supplementary Figure S2). For example, some patients had a CK P value greater than 5000 U/L. To give a reference point, male patients have a normal range of 30 - 200 U/L. However, since these outliers represent real clinical cases, they are retained in the training and test sets to allow the models to learn from extreme cases.

The eigenvalues of the principal components were analyzed to understand the data’s dimensionality. Around 90% of the explained variance was captured in the first few components. The most important features were Age, Gender, Status, ALT P, AST P, CK P, CRP P, Cl poc, Ery, and GGT P.

### Comparative Analysis across datasets

Table 3 shows the performance of each ML model using different datasets to classify PN and MY. To summarize the datasets during the experiments, Dataset A consists of 3,701 features, including demographics, laboratory test results, and ICD-10-GM codes. Dataset B is the baseline, consisting of age and gender. Dataset C contains age, gender, living status, and descriptive statistics of each laboratory test result. Dataset D only consists of deviations from the maximum and minimum, along with additional laboratory test results. Dataset E extends Dataset D by incorporating ICD-10-GM codes. Finally, PCA was performed on Dataset E and used the first 11 principal components, which resulted in Dataset F. Please refer to Table 1 for an overview of the datasets used in these experiments.

**Table 3.** Classification performance of our models on the test sets. Dataset A contains all the features. Dataset B consists of two features: age and gender. Dataset C consists of demographic and engineered features. Dataset D consists of demographic, engineered, and laboratory value features. Dataset E is similar to Dataset D but contains ICD-10-GM codes. Dataset F contains the top 11 principal components based on the dimensionality reduction of Dataset E using Principal Component Analysis. Upsampling is performed using SMOTE, while downsampling is performed randomly. LS stands for log-scaling, which refers to experiments done with log-scaling of non-normal features instead of normalization.

| Model | Accuracy | F1 Macro | AUC-ROC |
| --- | --- | --- | --- |
| Logistic Regression (Dataset B: Baseline) | 0.86 | 0.56 | 0.77 |
| Random Forest (Dataset B: Baseline) | 0.84 | 0.58 | 0.72 |
| Random Forest (Dataset A) | 0.85 | 0.65 | 0.80 |
| Random Forest (Dataset C) | 0.87 | 0.72 | 0.82 |
| Random Forest (Dataset D) | 0.91 | 0.78 | 0.90 |
| Random Forest (Dataset D & LS) | 0.91 | 0.78 | 0.90 |
| Random Forest (Dataset D: Upsampled) | 0.91 | 0.82 | 0.92 |
| Random Forest (Dataset D: Upsampled & LS) | 0.91 | 0.82 | 0.92 |
| XGBoost (Dataset D) | **0.93** | 0.84 | 0.91 |
| XGBoost (Dataset D: Upsampled) | **0.92** | **0.84** | 0.92 |
| XGBoost (Dataset D: Upsampled & LS) | **0.92** | **0.84** | 0.91 |
| Random Forest (Dataset E) | 0.91 | 0.79 | **0.93** |
| Random Forest (Dataset E: Downsampled) | 0.88 | 0.80 | 0.91 |
| Random Forest (Dataset E: Downsampled & LS) | 0.87 | 0.79 | 0.92 |
| Random Forest (Dataset E: Upsampled) | 0.91 | 0.82 | 0.92 |
| Random Forest (Dataset E: Upsampled & LS) | **0.92** | 0.83 | 0.92 |
| XGBoost (Dataset E) | **0.92** | 0.82 | 0.90 |
| XGBoost (Dataset E: Upsampled) | 0.91 | 0.81 | 0.92 |
| Random Forest (Dataset F: PCA) | 0.88 | 0.72 | 0.84 |

As shown in Table 3, Datasets D and E performed remarkably well; however, Datasets A, B, C, and F yielded less satisfactory results. In Dataset D, laboratory test results and demographic features (e.g., age and gender) appeared as the primary factors for distinguishing PN from MY. The addition of ICD-10-GM codes in Dataset E provided only a modest performance gain compared to Dataset D. Finally, when Dataset F is trained using the first 11 principal components on RF, this dataset performed worse compared to Datasets D and E.

Figure 7 shows the plots of the top-performing models for each dataset. See Supplementary Figure S5 for all results. The LR baseline and RF (Datasets A, C, and F) show similar performance when increasing the threshold (FPR). In particular, RF (Dataset C) distinguishes PN patients better at a lower threshold (FPR ≤ 0.2) until the RF (Dataset A and F) outperforms the model at a higher threshold (FPR> 0.2). The models trained on Datasets D and E showed strong performance (AUC-ROC ≥ 0.90).

**Fig. 6:**
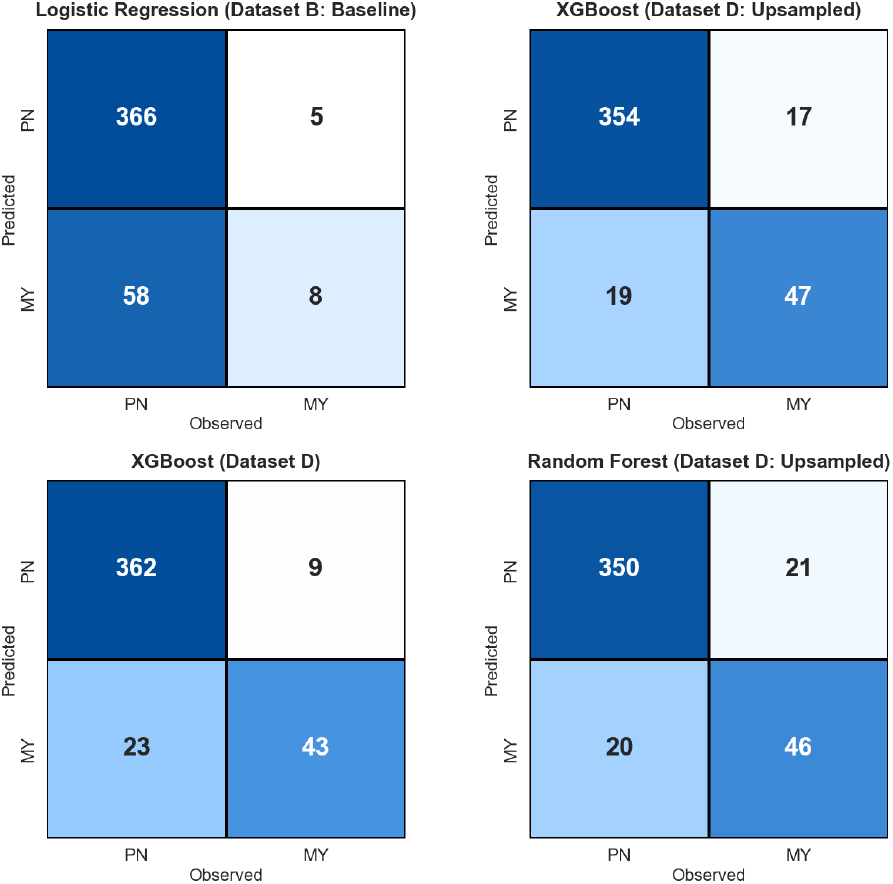
Confusion matrices of baseline and all the best performing models.

**Fig. 7:**
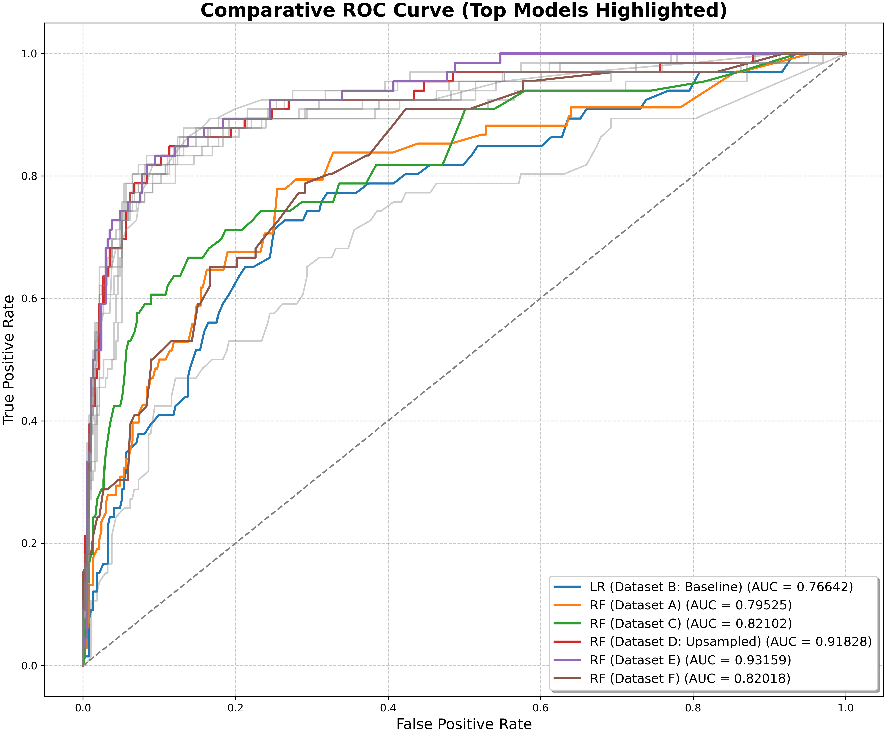
Comparative ROC Curve for top performing models for each dataset in the study. The plot also mentions the AUC scores of the models. LR stands for Logistic Regression; RF stands for Random Forest.

The AUC-PR scores offer another perspective when taking into account the class imbalance (See Figure 8). Although the AUC-PR score for PN remains high, the score for MY varies by model and dataset. In particular, the LR baseline and RF (Datasets A, C & F) struggle to classify MY. Similar to previous AUC-ROC scores, the models trained on Datasets D and E are capable of classifying most MY patients. For full results, refer to Supplementary Figure S6 and S7.

**Fig. 8:**
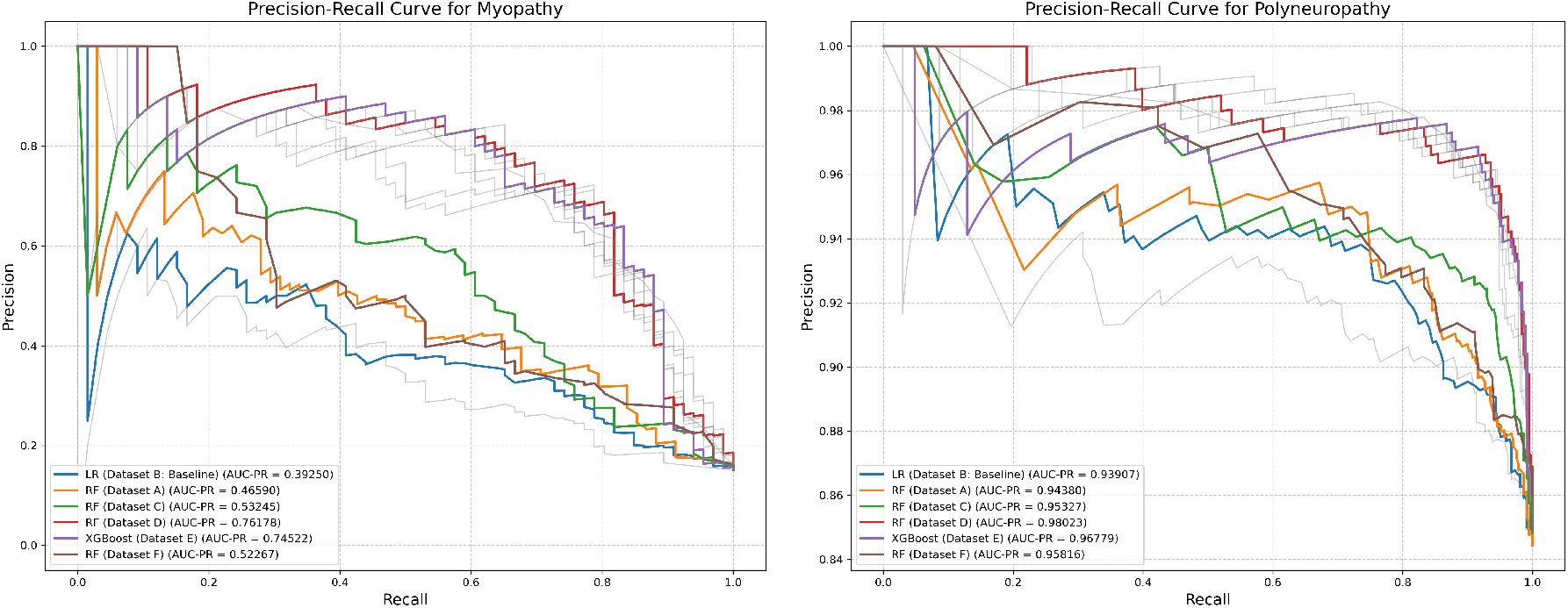
Precision-Recall Curve for Myopathy and Polyneuropathy of the top performing models for each dataset. LR stands for Logistic Regression; RF stands for Random Forest.

Figure 6 shows the best-performing datasets and models by F1 Macro, including the baseline for comparison. The LR (Dataset B) struggles to classify MY patients. An increase in true positive cases on MY classification occurs after adopting Dataset D. Notably, the upsampled datasets are accompanied by a considerable increase in true positive cases of MY patients at the cost of misclassifying PN patients. Supplementary Figure S1 shows the confusion matrices across all models and datasets.

### Principal Component Analysis visualization

To further explore Dataset E, dimensionality reduction is applied using PCA to investigate whether the diseases are well-separated. The 3D visualization (see Figure 5) and three pairwise scatter plots of the first three principal components (see Supplementary Figure S3) show in total 59% explained variance.

There exists a significant overlap between the PN and MY patients, consistent with the initial investigations on the raw dataset. This indicates, however, that given the features, it may not be possible to reduce the dimensionality further, as evidenced by the performance of Dataset F and the 3D and 2D scatter plots.

## Discussion

Machine learning (ML)–driven differentiation of polyneuropathy (PN) and myopathy (MY) on routine electronic health records (EHRs) represents an important translational step toward faster, data-supported pathways in neuromuscular medicine.

### Main findings

Clinically, our RF and XGBoost models achieved robust discrimination (F1 Macro 0.82-0.84; AUC-ROC 0.92-0.93), confirming that even moderately deep EHRs, when rigorously curated, contain sufficient signal to assist early separation of PN and MY in routine care settings. Methodologically, expert-driven feature curation added substantial diagnostic value over purely data-driven pipelines. Strategically, the German Forschungsdatenportal Gesundheit (FDPG) and its federated Data-Integration Centres (DICs) provide a scalable substrate for repeated model retraining and validation [54]. Looking forward, alignment with the emerging European Health Data Space (EHDS) promises cross-border expansion of neuromuscular ML tools and federated learning at a continental scale [55].

### Diagnostic Yield in Real-World Neurology

The achieved AUC-ROC (> 0.90) indicates clinically meaningful separation of PN and MY phenotypes despite overlapping symptomatology. For clinicians, this translates into a selection probability: a patient flagged by the model as MY with 0.9 posterior probability is roughly nine times more likely to harbour primary muscle pathology than peripheral nerve disease, enabling earlier electromyography or muscle biopsy referrals. Conversely, a high PN probability supports nerve-conduction studies and metabolic work-ups. Traditional diagnostic work-ups rely on stepwise exclusion that may span months and multiple specialties. Incorporating the ML flag at point-of-care could shorten this trajectory by highlighting atypical presentations (e.g. **Myofibrillar Myopathy**) that otherwise mimic PN, thereby reducing misclassification and inappropriate immunosuppression. In practice, integration of a simple dashboard within the hospital information system can provide a “second opinion” without replacing clinical judgment.

Previous studies have demonstrated the use of machine learning in EHR data for the recognition of rare diseases. For example, Cohen et al. [10] used a Support Vector Machine (SVM) to detect acute intermittent liver porphyria using a small and targeted dataset, while Garg et al. [45] achieved a high classification performance (F1 = 0.98) for cardiac amyloidosis using Random Forest in more structured clinical data. Lin et al. [56] combined Random Forest with linear discriminant analysis to classify neurodegenerative disorders based on blood biomarkers, although their work excluded routine clinical variables.

### Data Infrastructure: Current German Assets (Medizininformatik-Initiative and FDPG)

The Medizininformatik-Initiative (MII) has installed a network of DICs across all German university hospitals, exposing harmonised core datasets through a central FDPG portal [57]. A modular core dataset (in German “Kerndatensatz” (KDS)) encoded in HL7^®^ FHIR ensures syntactic and semantic consistency, while a national Broad Consent facilitates secondary use within strict data-protection safeguards. In 2024 alone, FDPG handled 19 registered research projects and 31 data-access requests, demonstrating operational readiness [58].

The German DIC network provides substantial advantages for neuromuscular machine learning applications through three key capabilities. The DIC network offers exceptional cohort scalability by enabling the pooling of more than 200, 000 neurology admissions annually across all German university hospitals, which allows for rapid accrual of rare neuromuscular cases that would otherwise be difficult to study at individual institutions [59, 60]. This large-scale data aggregation capability is particularly valuable for rare neuromuscular disorders where individual centers may see only a few cases per year.

The network facilitates powerful federated queries that allow researchers to execute distributed analytics across multiple hospitals without requiring the export of patient-level data, thereby preserving privacy while simultaneously increasing statistical power for research studies [61]. This federated approach enables multi-site analyses while maintaining strict data protection standards in compliance with German privacy regulations and the broader European Health Data Space framework.

Additionally, the system provides crucial longitudinal depth through the use of re-identifiable pseudonyms that enable comprehensive longitudinal tracking of disease progression and treatment response over time, which feeds sophisticated time-series machine learning models [62]. This capability is essential for understanding the natural history of neuromuscular diseases and developing predictive models that can inform clinical decision-making and patient management strategies.

### Path Toward a Learning Neuromuscular Health System

#### Clinical Decision Support Implementation

Successful clinical adoption demands transparent, explainable outputs. The recent AI Act requires high-risk AI systems to be designed so that the clinician and the patient can interpret the output. Clinicians who are unable to understand or interpret the output of the ML model will unlikely be able to state the information to the patient [63]. Therefore, we recommend measures such as Shapley values to provide “global” explanations that assess the overall logic behind the ML model and “local” explanations that reveal the reasons and risk factors for a patient classified as either PN or MY. These may include highlighting top-contributing lab anomalies (e.g., CK, AST), ICD-10-GM comorbidities (e.g., diabetes), and demographic modifiers (age, sex). Embedding these explanations in the electronic chart fosters clinician trust and peer review.

#### Expanding Machine-Learning Horizons

Future model iterations should exploit temporal trends and longitudinal patterns to enhance diagnostic precision. Current approaches aggregate laboratory values across time points, potentially losing critical trajectory information such as CK evolution patterns that may distinguish progressive myopathies from stable neuropathies. Advanced architectures including recurrent neural networks and temporal graph networks can capture these dynamic relationships, while irregular-time-series methods can accommodate the heterogeneous visit intervals characteristic of real-world neurology practice.

The federated infrastructure provided by EHDS and FDPG enables secure multi-institutional model development through distributed learning approaches [64]. Federated learning allows models to train across decentralized DIC nodes by exchanging model gradients rather than patient data, thereby mitigating data-silo bias while maintaining GDPR compliance—a fundamental prerequisite for EHDS participation [65].

Enhanced feature engineering should incorporate genetic and familial context, particularly given the hereditary nature of many neuromuscular disorders [66]. Family history, genetic testing results, and inheritance patterns could significantly improve model discrimination between acquired and inherited conditions [67]. Additionally, model performance depends critically on data quality and completeness. Standardized clinical documentation practices, consistent diagnostic coding, and systematic laboratory result recording will be essential to minimize missing values and ensure robust model generalisability across different clinical environments.

### Strengths and Limitations

Clinician-guided feature curation ensures biomedical plausibility by incorporating expert knowledge from UMG neuromuscular specialists, effectively bridging machine learning methodology with clinical expertise to identify diagnostically relevant biomarkers for polyneuropathy and myopathy differentiation. Rigorous evaluation metrics, including F1 Macro scores and Area Under the Curve Precision-Recall (AUC-PR), specifically address the class imbalance inherent to rare neuromuscular diseases, providing robust performance assessment that accounts for the 5.6:1 ratio of polyneuropathy to myopathy cases in our dataset.

A key limitation of this study is the lack of external validation. Although the models are trained and tested on the DIC dataset, it is currently unclear if the models are able to generalize to an entirely separate dataset in a different clinical environment and population distribution.

Despite the strong performance of Datasets D and E in differentiating PN and MY, it remains unclear whether ICD-10-GM codes could contribute to this outcome. The ICD-10-GM codes in Dataset E, mainly consist of symptoms and comorbidities not relevant to these diseases (see Supplementary Table S10). As disease-related symptoms are missing in the DIC dataset, ICD-10-GM codes contribute little to differentiating these diseases. Including both chronic complaints and acute symptoms are crucial in routine clinical practice. In addition, some patients have one or more pre-existing diseases, which affect the laboratory test results. These confounding factors could lead to deviations in laboratory test results and, therefore, may reflect on the results of classification on PN or MY.

## Vision Statement

By 2030, we envision an integrated learning health ecosystem where federated algorithms provide real-time diagnostic support, transforming lengthy diagnostic odysseys into precision medicine approaches that improve patient outcomes while advancing neuromuscular disease research through a harmonised European data infrastructure.

## Conclusion

Our work demonstrates that routine German EHRs, harmonised via MII-infrastructure, can serve as a robust foundation for developing clinically relevant ML models for PN-MY differentiation. Leveraging the upcoming EHDS to scale these models across institutions and borders will multiply their diagnostic reach, reduce inequities in orphan diseases, and accelerate the transition towards a new era of federated, patient-centred neuromuscular medicine. Continuous collaboration between clinicians, data scientists, patient advocates, and regulatory bodies is paramount to realise this vision while safeguarding patient autonomy and data security.

## Acknowledgement

We thank the Data Integration Center of the University Medical Center of Göttingen for providing access to the data used in this study.

## Author contributions

SA, NT, and VS conducted the computational analysis. JaZ, EN, and HW provided clinical expertise. JaZ and RR conceived, planned the study, oversaw the project, and secured the funding. JaZ and EN developed the ethical and legal workflow to obtain the data. All authors read, revised, and approved the final manuscript for submission.

## Ethical Considerations

This study was conducted following the Declaration of Helsinki and received approval from the ethics committees of the UMG (ref. 3/4/24, V1.0, 06.02.2024). The use of EHRs in this study adhered strictly to ethical standards and legal requirements. All patient data provided by the UMG DIC was de-identified and handled in compliance with the provisions of the Lower Saxony Data Protection Act (NDSG) and the EU General Data Protection Regulation (EU GDPR) to ensure confidentiality and privacy. Given the sensitive nature of EHR data, particular emphasis was placed on maintaining patient autonomy and minimizing risks of re-identification. The study also followed established protocols for secure data storage and access control. Access to the data was only possible through a managed access procedure that included user ID and password protection, automatic screen lock, virus protection, a firewall, hardware encryption, and a key-locking system. Consequently, only authorized persons had access rights. Collaboration with UMG data protection officers and ethical boards was maintained to uphold the highest standards of ethical governance.

## Data Availability

The code used in this study is available at https://gitlab.sdu.dk/screen4care/classifying-pn-and-my. The dataset is not publicly available due to privacy restrictions but may be available from the corresponding author upon reasonable request and with permission from the Medical Data Integration Center of the University of Göttingen.

