## Supplementary data (Table S1-S11; Figure S1-S7) for "Classifying polyneuropathy and myopathy patients on Electronic Health Records"

### ICD-10GM codes

We present four tables showing an overview of ICD-10GM codes for patients diagnosed with polyneuropathy and myopathy. Table 1 and 2 represent 2,494 unique patients from the raw dataset. This includes patients who are diagnosed with both diseases and patients who are not diagnosed. The ICD-10GM codes are taken from gesund.bund.de [ges].

Table 3 and 4 represent all patients from Dataset A. Here, we filtered out patients not meeting the following criteria: 1. Patients are diagnosed with only one disease that belongs to polyneuropathy or myopathy. 2. The patients did not do any laboratory tests. This results in 2,181 unique patients. It is important to note that patients may have multiple ICD-10 codes associated with either polyneuropathy or myopathy.

| **ICD Code** | **Name** | **ICD Code Frequency** | **Percentage** |
|---|---|---|---|
| G62.88 | Other specified polyneuropathies | 1200 | 39.22 |
| G62.9 | Polyneuropathy, unspecified | 477 | 15.59 |
| G61.8 | Other inflammatory polyneuropathies | 422 | 13.79 |
| G62.80 | Critical illness polyneuropathy | 271 | 8.86 |
| G72.80 | Critical illness myopathy | 182 | 5.95 |
| G62.0 | Drug-induced polyneuropathy | 119 | 3.89 |
| G61.0 | Guillain-Barré syndrome | 98 | 3.20 |
| G63.2 | Diabetic polyneuropathy | 68 | 2.22 |
| G60.0 | Hereditary motor and sensory neuropathy | 48 | 1.57 |
| G62.1 | Alcoholic polyneuropathy | 42 | 1.37 |
| G61.9 | Inflammatory polyneuropathy, unspecified | 24 | 0.78 |
| G72.88 | Other specified myopathies | 15 | 0.49 |
| G60.8 | Other hereditary and idiopathic neuropathies | 13 | 0.42 |
| G70.0 | Myasthenia gravis | 11 | 0.36 |
| G62.2 | Polyneuropathy due to other toxic agents | 9 | 0.29 |
| G72.4 | Inflammatory myopathy, not elsewhere classified | 8 | 0.26 |
| G72.9 | Myopathy, unspecified | 8 | 0.26 |
| G60.9 | Hereditary and idiopathic neuropathy, unspecified | 6 | 0.20 |
| G63.4 | Polyneuropathy in nutritional deficiency | 5 | 0.16 |
| G71.0 | Muscular dystrophy | 4 | 0.13 |
| G60.3 | Idiopathic progressive neuropathy | 4 | 0.13 |
| G63.5 | Polyneuropathy in systemic connective tissue disorders | 4 | 0.13 |
| G73.7 | Myopathy in other diseases classified elsewhere | 3 | 0.10 |
| G71.1 | Myotonic disorders | 3 | 0.10 |
| G61.1 | Serum neuropathy | 2 | 0.07 |
| G60.1 | Refsum disease | 2 | 0.07 |
| G71.9 | Primary disorder of muscle, unspecified | 2 | 0.07 |
| G64 | Other disorders of peripheral nervous system | 2 | 0.07 |
| G63.8 | Polyneuropathy in other diseases classified elsewhere | 1 | 0.03 |
| G60.2 | Neuropathy in association with hereditary ataxia | 1 | 0.03 |
| G63.0 | Polyneuropathy in infectious and parasitic diseases classified elsewhere | 1 | 0.03 |
| G63.6 | Polyneuropathy in other musculoskeletal disorders | 1 | 0.03 |
| G70.8 | Other specified myoneural disorders | 1 | 0.03 |
| G71.3 | Mitochondrial myopathy, not elsewhere classified | 1 | 0.03 |
| G73.5 | Myopathy in endocrine diseases | 1 | 0.03 |
| G73.0 | Myasthenic syndromes in endocrine diseases | 1 | 0.03 |

Table 1: Frequency and Percentage of ICD-10GM Codes for Polyneuropathy. ICD-10GM codes for Myopathy also exist since the patient can suffer from both diseases. The percentage is calculated by the frequency divided by the total sum of frequencies.

| ICD Code | Name | ICD Code Frequency | Percentage |
|---|---|---|---|
| G72.80 | Critical illness myopathy | 188 | 32.70 |
| G62.80 | Critical illness polyneuropathy | 181 | 31.48 |
| G72.88 | Other specified myopathies | 162 | 28.17 |
| G71.0 | Muscular dystrophy | 107 | 18.61 |
| G72.4 | Inflammatory myopathy, not elsewhere classified | 90 | 15.65 |
| G71.1 | Myotonic disorders | 77 | 13.39 |
| G72.9 | Myopathy, unspecified | 75 | 13.04 |
| G62.88 | Other specified polyneuropathies | 39 | 6.78 |
| G71.8 | Other primary disorders of muscles | 28 | 4.87 |
| G70.0 | Myasthenia gravis | 14 | 2.43 |
| G61.8 | Other inflammatory polyneuropathies | 11 | 1.91 |
| G62.9 | Polyneuropathy, unspecified | 11 | 1.91 |
| G63.2 | Diabetic polyneuropathy | 10 | 1.74 |
| G72.0 | Drug-induced myopathy | 6 | 1.04 |
| G71.2 | Congenital myopathies | 6 | 1.04 |
| G72.3 | Periodic paralysis | 5 | 0.87 |
| G71.3 | Mitochondrial myopathy, not elsewhere classified | 4 | 0.70 |
| G71.9 | Primary disorder of muscle, unspecified | 4 | 0.70 |
| G70.8 | Other specified myoneural disorders | 4 | 0.70 |
| G73.7 | Myopathy in other diseases classified elsewhere | 3 | 0.52 |
| G62.0 | Drug-induced polyneuropathy | 1 | 0.17 |
| G73.6 | Myopathy in metabolic diseases | 1 | 0.17 |
| G73.5 | Myopathy in endocrine diseases | 1 | 0.17 |
| G73.0 | Myasthenic syndromes in endocrine diseases | 1 | 0.17 |
| G72.1 | Alcoholic myopathy | 1 | 0.17 |
| G61.1 | Serum neuropathy | 1 | 0.17 |
| G70.9 | Myoneural disorder, unspecified | 1 | 0.17 |
| G60.0 | Hereditary motor and sensory neuropathy | 1 | 0.17 |
| G63.4 | Polyneuropathy in nutritional deficiency | 1 | 0.17 |
| G61.0 | Guillain-Barré syndrome | 1 | 0.17 |

Table 2: Frequency and Percentage of ICD-10GM Codes for Myopathy. ICD-10GM codes for Polyneuropathy also exist since the patient can suffer from both diseases. The percentage is calculated by the frequency divided by the total sum of frequencies.

| ICD-10GM Code | Name | Frequency | Percentage |
|---|---|---|---|
| G62.88 | Other specified polyneuropathies | 1149 | 45.69 |
| G62.9 | Polyneuropathy, unspecified | 444 | 17.65 |
| G61.8 | Other inflammatory polyneuropathies | 404 | 16.06 |
| G62.0 | Drug-induced polyneuropathy | 117 | 4.65 |
| G61.0 | Guillain-Barré syndrome | 95 | 3.78 |
| G62.80 | Critical illness polyneuropathy | 90 | 3.58 |
| G63.2 | Diabetic polyneuropathy | 58 | 2.31 |
| G60.0 | Hereditary motor and sensory neuropathy | 46 | 1.83 |
| G62.1 | Alcoholic polyneuropathy | 42 | 1.67 |
| G61.9 | Inflammatory polyneuropathy, unspecified | 24 | 0.95 |
| G60.8 | Other hereditary and idiopathic neuropathies | 13 | 0.52 |
| G62.2 | Polyneuropathy due to other toxic agents | 8 | 0.32 |
| G60.9 | Hereditary and idiopathic neuropathy, unspecified | 6 | 0.24 |
| G60.3 | Idiopathic progressive neuropathy | 4 | 0.16 |
| G63.4 | Polyneuropathy in nutritional deficiency | 4 | 0.16 |
| G63.5 | Polyneuropathy in systemic connective tissue disorders | 4 | 0.16 |
| G64 | Other disorders of peripheral nervous system | 2 | 0.08 |
| G60.2 | Neuropathy in association with hereditary ataxia | 1 | 0.04 |
| G61.1 | Serum neuropathy | 1 | 0.04 |
| G63.0 | Polyneuropathy in infectious and parasitic diseases classified elsewhere | 1 | 0.04 |
| G63.6 | Polyneuropathy in other musculoskeletal disorders | 1 | 0.04 |
| G63.8 | Polyneuropathy in other diseases classified elsewhere | 1 | 0.04 |

Table 3: Frequency and Percentage of ICD-10GM Codes for Polyneuropathy (n = 2515). The percentage is calculated by the frequency divided by the total sum of frequencies.

| ICD-10GM Code | Name | Frequency | Percentage |
|---|---|---|---|
| G72.88 | Other specified myopathies | 146 | 28.80 |
| G71.0 | Muscular dystrophy | 89 | 17.55 |
| G72.4 | Inflammatory myopathy, not elsewhere classified | 79 | 15.58 |
| G72.9 | Myopathy, unspecified | 65 | 12.82 |
| G71.1 | Myotonic disorders | 63 | 12.43 |
| G71.8 | Other primary disorders of muscles | 28 | 5.52 |
| G72.80 | Critical illness myopathy | 6 | 1.18 |
| G72.0 | Drug-induced myopathy | 6 | 1.18 |
| G71.2 | Congenital myopathies | 6 | 1.18 |
| G72.3 | Periodic paralysis | 5 | 0.99 |
| G70.8 | Other specified myoneural disorders | 3 | 0.59 |
| G71.3 | Mitochondrial myopathy, not elsewhere classified | 3 | 0.59 |
| G70.0 | Myasthenia gravis | 3 | 0.59 |
| G71.9 | Primary disorder of muscle, unspecified | 2 | 0.39 |
| G70.9 | Myoneural disorder, unspecified | 1 | 0.20 |
| G72.1 | Alcoholic myopathy | 1 | 0.20 |
| G73.6 | Myopathy in metabolic diseases | 1 | 0.20 |

Table 4: Frequency and Percentage of ICD-10GM Codes for Myopathy (n = 507). The percentage is calculated by the frequency divided by the total sum of frequencies.

#### ICD-10GM - Examinations, symptoms and comorbidities

Table 5 and 6 show the most frequent ICD-10GM codes that occurred at least once for polyneuropathy and myopathy. Diagnosis codes for diagnosing polyneuropathy and myopathy are excluded from the tables.

| **ICD-10GM Code** | **Name** | **Percentage** |
|---|---|---|
| Z11 | Special screening examination for infectious and parasitic diseases | 66.09 |
| U99.0 | Special screening examination for SARS-CoV-2 | 61.76 |
| I10.00 | Benign essential hypertension<br>Without mention of hypertensive urgency | 50.69 |
| E87.6 | Hypokalaemia | 17.09 |
| E78.5 | Hyperlipidaemia, unspecified | 15.77 |
| Z92.2 | Personal history of long-term (current) use of other medicaments | 14.98 |
| N39.0 | Urinary tract infection, site not specified | 12.97 |
| I25.13 | Atherosclerotic heart disease: Triple-vessel coronary artery disease | 12.39 |
| E11.90 | Type 2 diabetes mellitus Without complications Controlled | 12.29 |
| D90 | Immune compromise due to radiation, chemotherapy or other immunosuppressive measures | 12.08 |
| E03.8 | Other specified hypothyroidism | 11.87 |
| J96.00 | Acute respiratory failure, not elsewhere classified Type 1 [with hypoxia] | 11.60 |
| B96.2 | Escherichia coli [E. coli] as the cause of diseases classified to other chapters | 11.55 |
| I48.0 | Paroxysmal atrial fibrillation | 11.55 |
| Z92.1 | Personal history of long-term (current) use of anticoagulants | 11.02 |

Table 5: Top 15 most frequent ICD-10GM codes for polyneuropathy (n = 1853). ICD-10GM codes for diagnosing polyneuropathy are excluded here.

| **ICD-10GM Code** | **Name** | **Percentage** |
|---|---|---|
| Z11 | Special screening examination for infectious and parasitic diseases | 53.14 |
| U99.0 | Special screening examination for SARS-CoV-2 | 52.86 |
| I10.00 | Benign essential hypertension<br>Without mention of hypertensive urgency | 21.14 |
| E11.90 | Type 2 diabetes mellitus Without complications Controlled | 9.71 |
| I48.0 | Paroxysmal atrial fibrillation | 7.71 |
| D90 | Immune compromise due to radiation, chemotherapy or other immunosuppressive measures | 6.29 |
| M79.10 | Myalgia Multiple sites | 6.0 |
| E03.8 | Other specified hypothyroidism | 6.0 |
| E87.6 | Hypokalaemia | 4.86 |
| E78.5 | Hyperlipidaemia, unspecified | 4.57 |
| R13.9 | Other and unspecified dysphagia | 4.29 |
| M60.80 | Other myositis Multiple sites | 4.0 |
| M62.80 | Other specified disorders of muscle Multiple sites | 3.71 |
| N39.0 | Urinary tract infection, site not specified | 3.43 |
| E06.3 | Autoimmune thyroiditis | 3.14 |

Table 6: Top 15 most frequent ICD-10GM codes for myopathy (n = 328). ICD-10GM codes for diagnosing myopathy are excluded here.

### Features of Dataset B, C, D, and E

| **Feature** | **Data type** |
|---|---|
| Age | Numeric |
| Gender | Binary |

Table 7: Dataset B containing age and gender as features. We consider this dataset as a baseline.

| **Feature** | **Data type** |
|---|---|
| mean_CRP P | Numeric |
| mean_Cl poc | Numeric |
| mean_Glu POC o | Numeric |
| mean_Glu po | Numeric |
| mean_K poct | Numeric |
| mean_Krea P | Numeric |
| mean_La poc | Numeric |
| mean_LaPoA | Numeric |
| mean_Na poc | Numeric |
| mean_iCa po | Numeric |
| std_CRP P | Numeric |
| std_Cl poc | Numeric |
| std_Glu POC o | Numeric |
| std_Glu po | Numeric |
| std_K poct | Numeric |
| std_Krea P | Numeric |
| std_La poc | Numeric |
| std_LaPoA | Numeric |
| std_Na poc | Numeric |
| std_iCa po | Numeric |
| count_CRP P | Numeric |
| count_Cl poc | Numeric |
| count_Glu POC o | Numeric |
| count_Glu po | Numeric |
| count_K poct | Numeric |
| count_Krea P | Numeric |
| count_La poc | Numeric |
| count_LaPoA | Numeric |
| count_Na poc | Numeric |
| count_iCa po | Numeric |
| min_CRP P | Numeric |
| min_Cl poc | Numeric |
| min_Glu POC o | Numeric |
| min_Glu po | Numeric |
| min_K poct | Numeric |
| min_Krea P | Numeric |
| min_La poc | Numeric |
| min_LaPoA | Numeric |
| min_Na poc | Numeric |
| min_iCa po | Numeric |
| max_CRP P | Numeric |
| max_Cl poc | Numeric |
| max_Glu POC o | Numeric |
| max_Glu po | Numeric |
| max_K poct | Numeric |
| max_Krea P | Numeric |
| max_La poc | Numeric |
| max_LaPoA | Numeric |
| max_Na poc | Numeric |
| max_iCa po | Numeric |

Table 8: Dataset C containing demographics and descriptive statistics of each laboratory test result (continued on next page).

| Feature | Data type |
|---|---|
| last_CRP P | Numeric |
| last_Cl poc | Numeric |
| last_Glu POC o | Numeric |
| last_Glu po | Numeric |
| last_K poct | Numeric |
| last_Krea P | Numeric |
| last_La poc | Numeric |
| last_LaPoA | Numeric |
| last_Na poc | Numeric |
| last_iCa po | Numeric |
| CRP P_abnormal_rate | Numeric |
| Cl poc_abnormal_rate | Numeric |
| Glu POC o_abnormal_rate | Numeric |
| Glu po_abnormal_rate | Numeric |
| K poct_abnormal_rate | Numeric |
| Krea P_abnormal_rate | Numeric |
| La poc_abnormal_rate | Numeric |
| LaPoA_abnormal_rate | Numeric |
| Na poc_abnormal_rate | Numeric |
| iCa po_abnormal_rate | Numeric |
| Age | Numeric |
| Gender | Binary |
| Status | Binary |

Table 8: Dataset C containing demographics and descriptive statistics of each laboratory test result (continued).

| **Feature** | **Data type** |
|---|---|
| Age | Numeric |
| Gender | Binary |
| Status | Binary |
| ALT P | Numeric |
| AST P | Numeric |
| CK P | Numeric |
| CRP P | Numeric |
| Cl poc | Numeric |
| Ery | Numeric |
| GGT P | Numeric |
| Glu POC o | Numeric |
| Glu po | Numeric |
| Gluc-P | Numeric |
| HB | Numeric |
| HBA1cM | Numeric |
| HK | Numeric |
| HbA1c | Numeric |
| IgA S | Numeric |
| IgG S | Numeric |
| IgM S | Numeric |
| K P | Numeric |
| K poct | Numeric |
| Krea P | Numeric |
| Ku | Numeric |
| La poc | Numeric |
| LaPoA | Numeric |
| Lac P | Numeric |
| Leuko | Numeric |
| MCH | Numeric |
| MCHC | Numeric |
| MCV | Numeric |
| Mi-2 alpha | Numeric |
| Mi-2 beta | Numeric |
| Na P | Numeric |
| Na poc | Numeric |
| Quick | Numeric |
| THR | Numeric |
| TIF1 gamma | Numeric |
| TSH P | Numeric |
| TSH rP | Numeric |
| aPTT | Numeric |
| eGFR ab 18 | Numeric |
| iCa po | Numeric |
| Deviation_from_Min | Numeric |
| Deviation_from_Max | Numeric |
| Days_Stayed | Numeric |

Table 9: Dataset D containing patient demographics, laboratory tests, and engineered data used as features.

| Feature | Data type |
|---|---|
| Age | Numeric |
| Gender | Binary |
| Status | Binary |
| ALT P | Numeric |
| AST P | Numeric |
| CK P | Numeric |
| CRP P | Numeric |
| Cl poc | Numeric |
| Ery | Numeric |
| GGT P | Numeric |
| Glu POC o | Numeric |
| Glu po | Numeric |
| Gluc-P | Numeric |
| HB | Numeric |
| HBA1cM | Numeric |
| HK | Numeric |
| HbA1c | Numeric |
| IgA S | Numeric |
| IgG S | Numeric |
| IgM S | Numeric |
| K P | Numeric |
| K poct | Numeric |
| Krea P | Numeric |
| Ku | Numeric |
| La poc | Numeric |
| LaPoA | Numeric |
| Lac P | Numeric |
| Leuko | Numeric |
| MCH | Numeric |
| MCHC | Numeric |
| MCV | Numeric |
| Mi-2 alpha | Numeric |
| Mi-2 beta | Numeric |
| Na P | Numeric |
| Na poc | Numeric |
| Quick | Numeric |
| THR | Numeric |
| TIF1 gamma | Numeric |
| TSH P | Numeric |
| TSH rP | Numeric |
| aPTT | Numeric |
| eGFR ab 18 | Numeric |
| iCa po | Numeric |
| ICD_B96.2 | Binary |
| ICD_D62 | Binary |
| ICD_D90 | Binary |
| ICD_E03.8 | Binary |
| ICD_E06.3 | Binary |
| ICD_E11.90 | Binary |
| ICD_E53.8 | Binary |
| ICD_E78.5 | Binary |
| ICD_E87.6 | Binary |
| ICD_G82.33 | Binary |
| ICD_I10.00 | Binary |

Table 10: Dataset E containing patient demographics, laboratory tests, ICD-10GM codes, and engineered data used as features (continued).

| Feature | Data type |
|---|---|
| ICD_I25.13 | Binary |
| ICD_I48.0 | Binary |
| ICD_J45.9 | Binary |
| ICD_J96.00 | Binary |
| ICD_M33.2 | Binary |
| ICD_M60.80 | Binary |
| ICD_M60.85 | Binary |
| ICD_M60.86 | Binary |
| ICD_M62.80 | Binary |
| ICD_M79.10 | Binary |
| ICD_M79.18 | Binary |
| ICD_M81.98 | Binary |
| ICD_N39.0 | Binary |
| ICD_R13.9 | Binary |
| ICD_R20.1 | Binary |
| ICD_R20.2 | Binary |
| ICD_R26.8 | Binary |
| ICD_Z92.1 | Binary |
| ICD_Z92.2 | Binary |
| Deviation_from_Min | Numeric |
| Deviation_from_Max | Numeric |
| Days_Stayed | Numeric |

Table 10: Dataset E containing patient demographics, laboratory tests, ICD-10GM codes, and engineered data used as features.

### Grid Search - Hyperparameters

| Models | Hyperparameters |
|---|---|
| Random Forest | n_estimators = [50, 100, 200]<br>max_depth = [None, 10, 20, 30]<br>min_samples_split = [2, 5, 10]<br>min_samples_leaf = [1, 2, 4] |
| XGBoost | n_estimators = [50, 100, 200]<br>learning_rate = [0.01, 0.1, 0.2]<br>max_depth = [3, 5, 7]<br>subsample = [0.7, 0.8, 1.0]<br>colsample_bytree = [0.7, 0.8, 1.0]<br>gamma = [0, 0.1, 0.2] |

Table 11: List of hyperparameters for Grid Search hyperparameter optimization.

### Supplementary Results

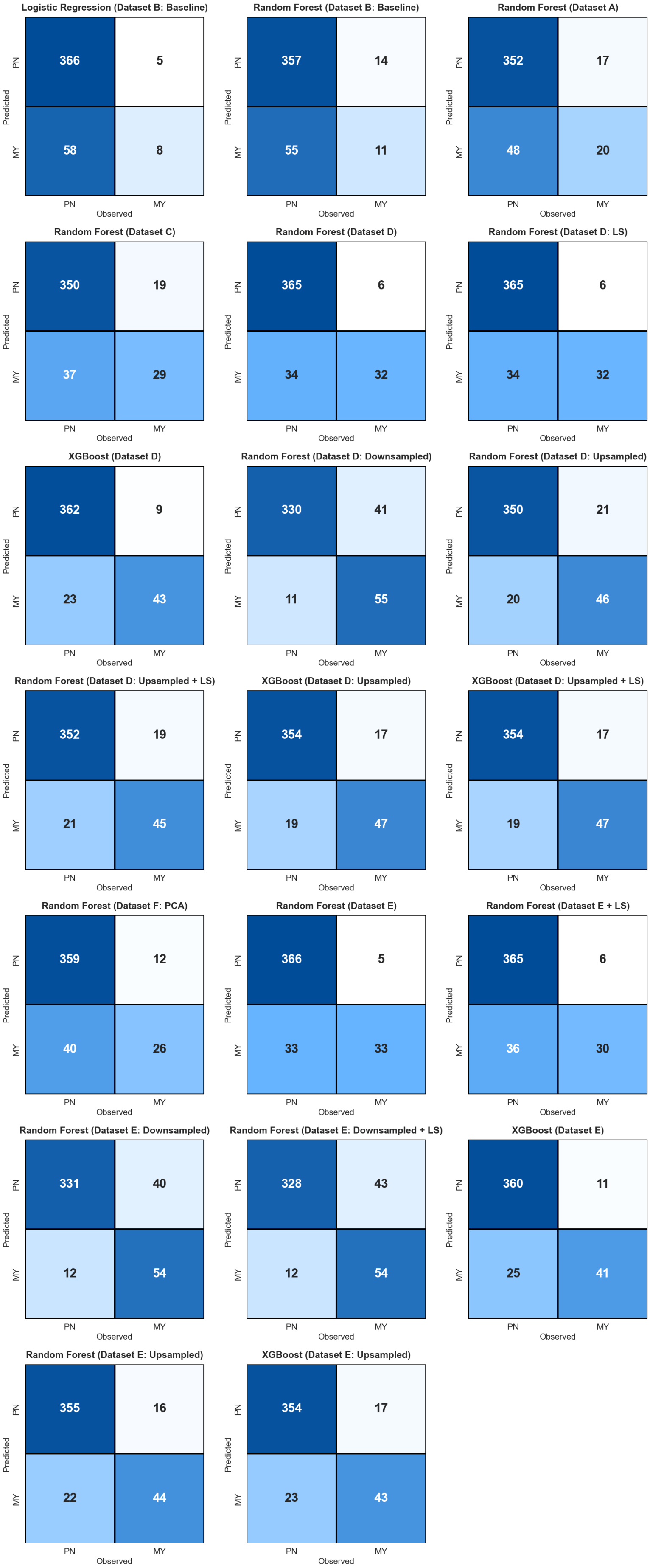


Figure 1: Confusion matrices of different datasets and Machine Learning models.

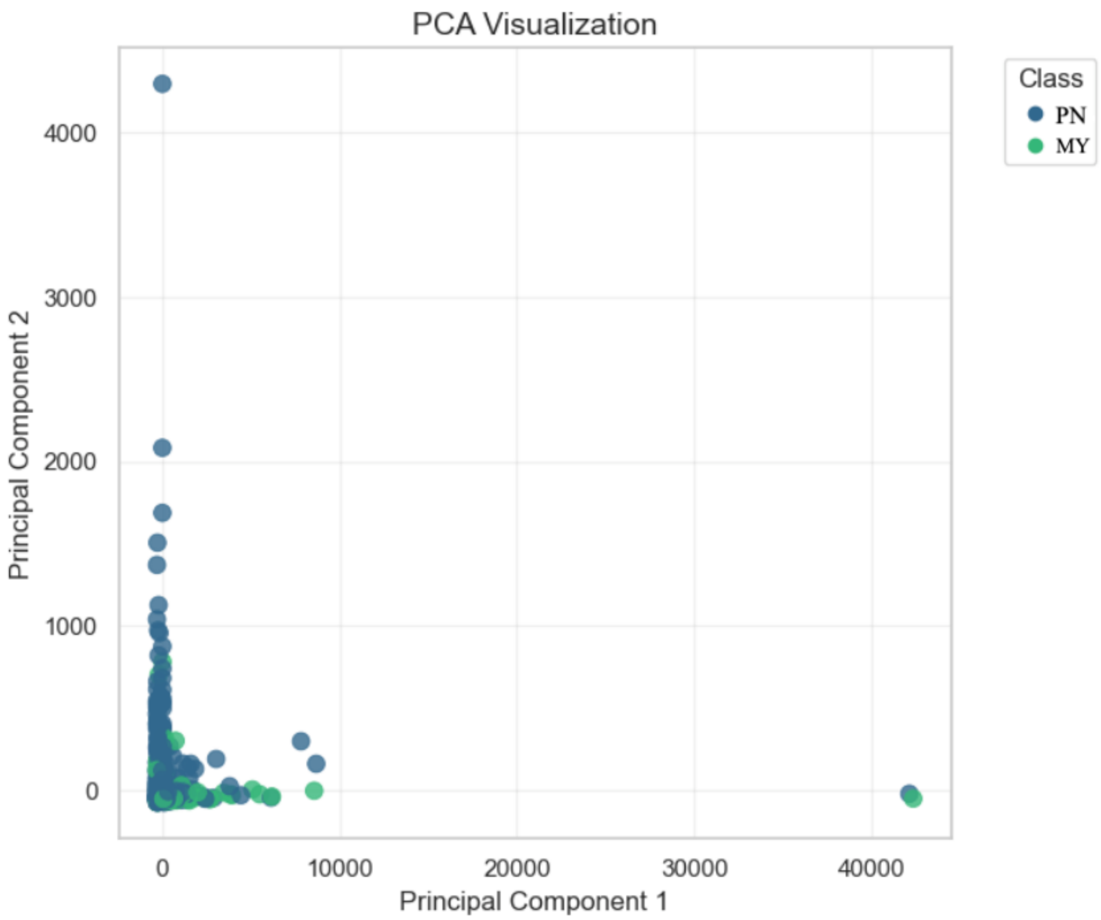


Figure 2: PCA plot on Dataset A. PN stands for polyneuropathy and MY for myopathy.

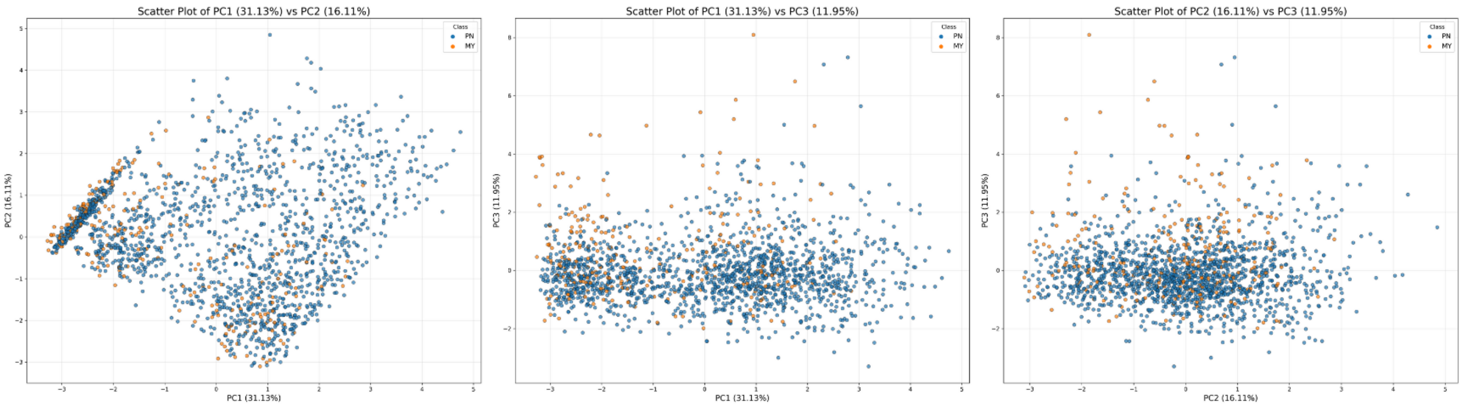


Figure 3: PCA scatter plot on Dataset E. PN stands for polyneuropathy and MY for myopathy.

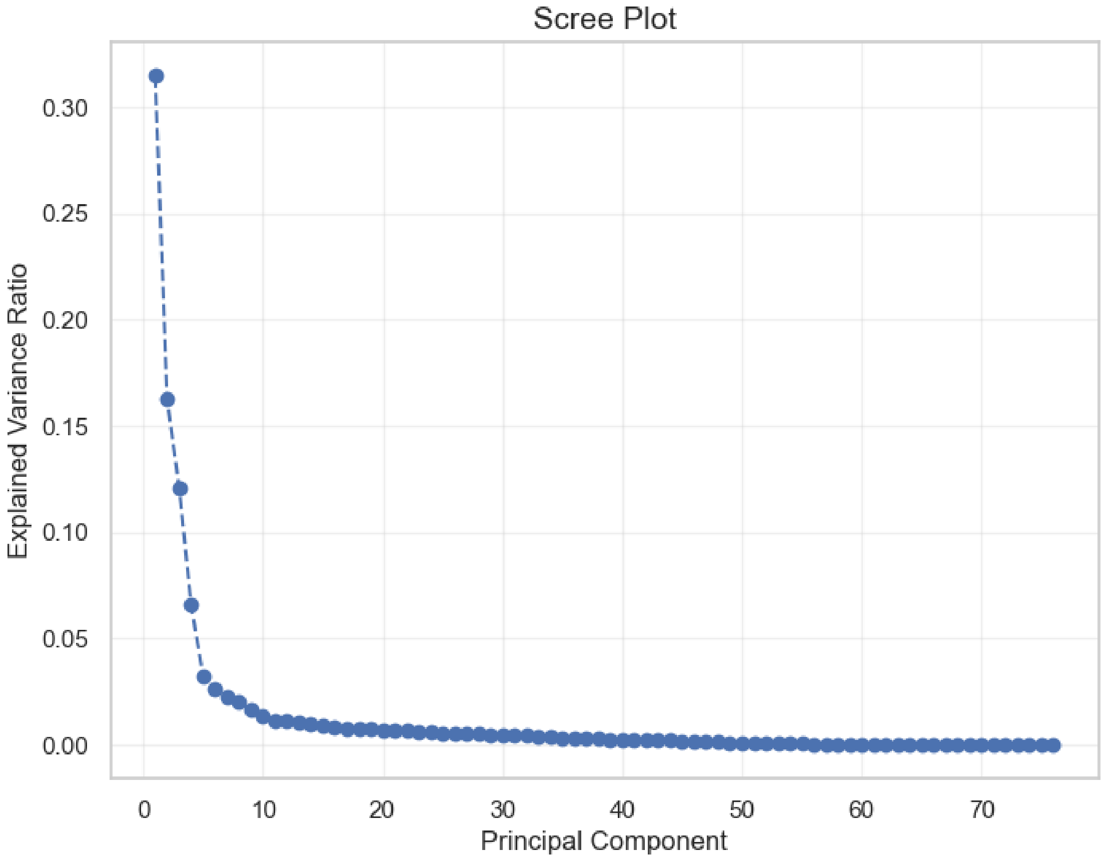


Figure 4: After performing dimensionality reduction on Dataset E, we created a Scree plot for deciding which first $n$ principal components we decided to choose for forming Dataset F. We chose the first 11 principal components for training our Random Forest model.

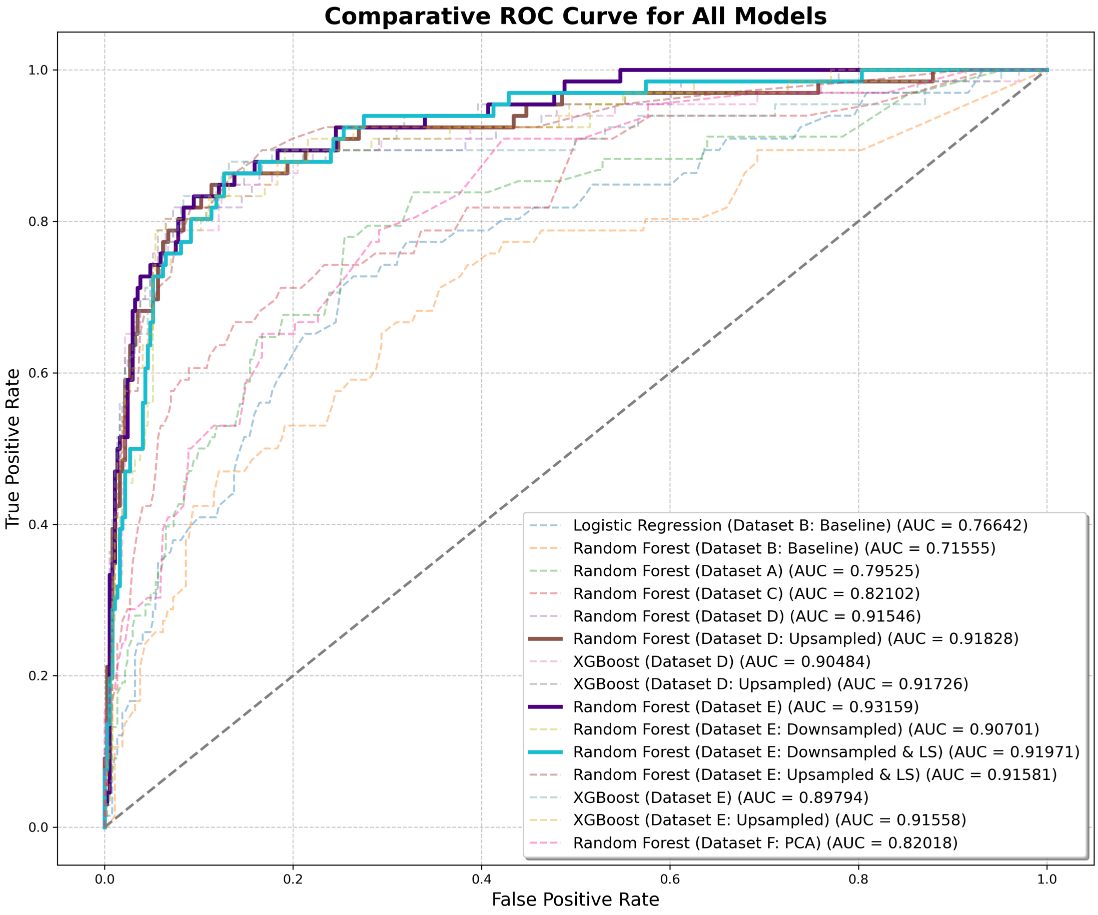


Figure 5: ROC curve showing results of all models. The top three models are highlighted.

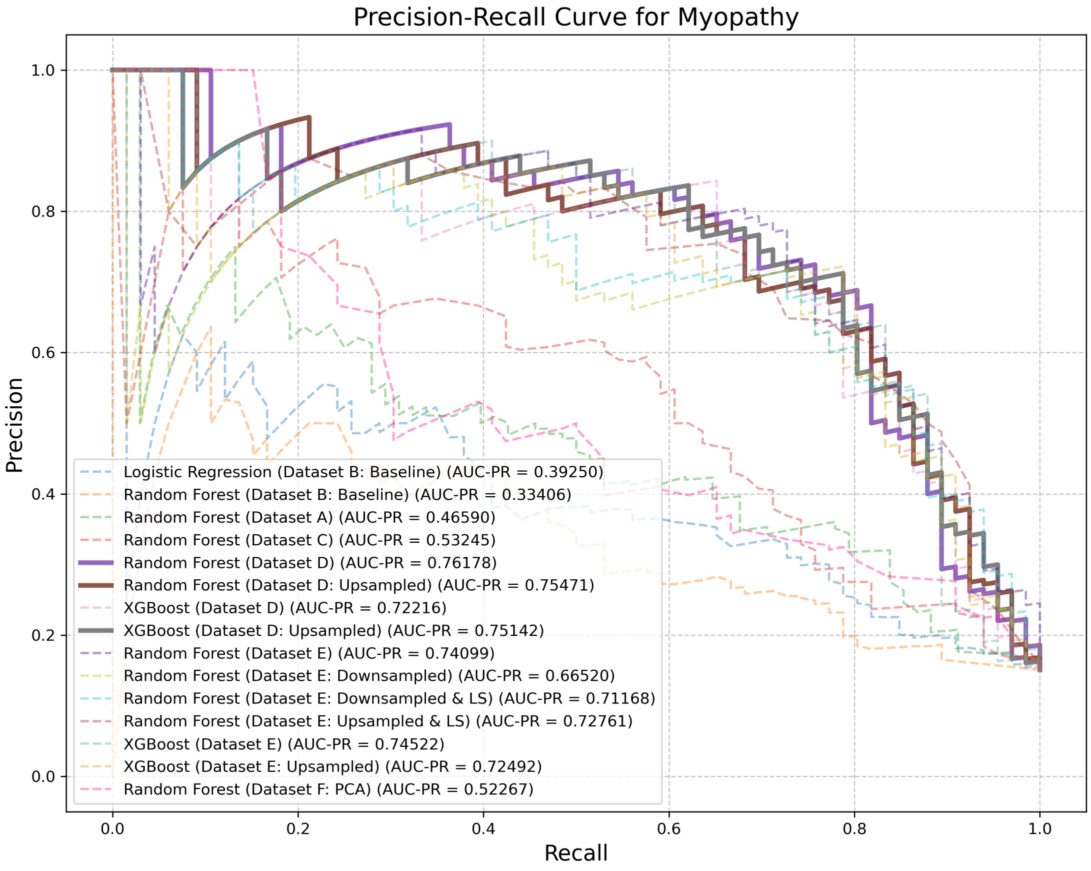


Figure 6: PR curve for myopathy showing results of all models. The top three models are highlighted.

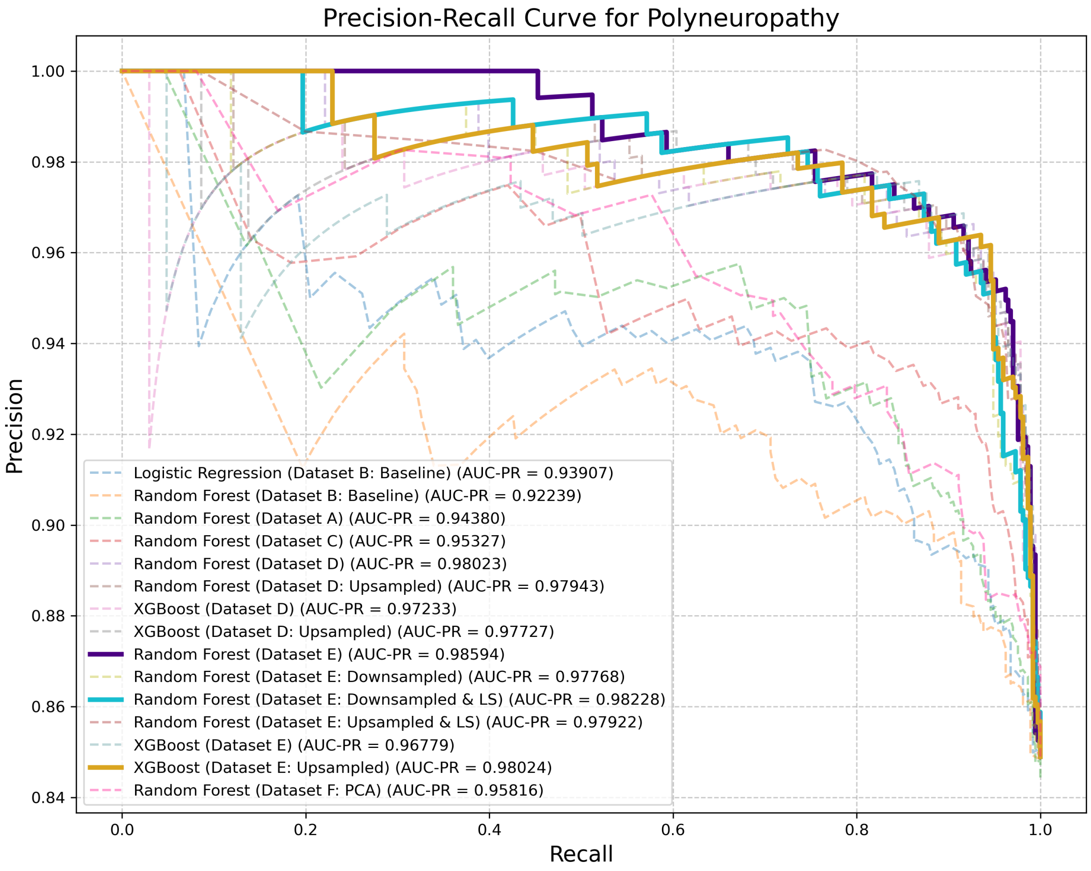


Figure 7: PR curve for polyneuropathy showing results of all models. The top three models are highlighted.
